# Telehealth during and beyond the COVID-19 Pandemic: Evidence from Licensed Dietitians in an Emerging Economy

**DOI:** 10.1101/2024.09.19.24314030

**Authors:** Maya Assaad, Nour Chamma, Miroslav Mateev, Rana Rizk

## Abstract

**Background:** The SARS-Cov-2 pandemic (COVID-19) sudden onset disrupted the direct access to face-to-face nutrition consultation fostering the rapid adoption of Telehealth by dietitians.

**Objective:** This study investigates Telehealth use among Lebanese Licensed Dietitians (LDs) amid COVID-19, in the absence of national Telehealth practical guidelines (TPG), and offers insights on Telehealth application under uncertainties of pandemic event, economic crisis, and destructed infrastructure occurring simultaneously in Lebanon.

**Design:** A cross-sectional study during March 2023, using an anonymous online survey (44 questions), diffused through the Lebanese Order of Dietitians and social media platforms.

**Participants:** The sample (n=94) consisted mostly of females (98.9%) and young dietitians (mean(SD) age: 30.54(6.41) years) having a mean(SD) of 7.89(5.7) years of experience. Most respondents identified clinical nutrition as their primary practice area (87.2%), mostly in weight management (84%).

**Main outcome measures:** Telehealth experience amid COVID-19 including tools utilized in remote consultations, barriers, facilitators, and perspectives of Telehealth use.

**Statistical analyses:** Descriptive analysis (counts, frequencies) using SPSS version 28.

**Results:** Although 48.4% of LDs reported using Telehealth prior COVID-19, this proportion increased to 97.8% during the pandemic. The most common tools used were WhatsApp (90.3%), Zoom (72.0%), and E-mails (41.9%). Reported barriers included bad internet connection (74.2%), patients preferring face-to-face consultation (61.3%), and patients lacking technical literacy (33.3%); benefits included scheduling and time flexibility (83.9%), decrease in practice-related costs (77.4%), and compliance with social distancing measures (53.8%). The majority agreed that Telehealth is needed (78.5%) and applicable in the Lebanese context (64.6%) and expressed the need for Telehealth trainings (78.5%) and TPG for nutrition care (74.2%).

**Conclusions:** This study recognizes increasing use of Telehealth in Lebanon, warranting the development of Telehealth nutrition care infrastructure comprising national regulations and evidence-based practical guidelines to respond to the innovation in the healthcare industry, and to assure Telehealth sustainability in LDs’ routine practice.

## Introduction

The SARS-Cov-2 pandemic (COVID-19) sudden onset caused severe lifestyle changes impacting negatively on the mental and physical health of individuals. Moreover, it severely disrupted the access to hospitals and clinics for face-to-face consultation and therapy. To overcome these challenges, a paradigm shift toward digital health and remote clinical services was seen among healthcare providers and dietitians.^1^ Facilitated by the emergence of information and communication technology (ICT) and direct-to-consumer virtual devices for data collection, Telehealth became an alternative and effective tool for nutrition care in time of pandemic.^2,3,4^

The public health emergency declaration of COVID-19 by the World Health Organization (WHO) followed by the lockdown imposed in March 2020 in Lebanon, have similarly exposed the medical sector to remote consultations, which became the “significant mediator for the resilience of the care system”.^5^ Prior to COVID-19, the country was in the primary phase of incorporating Telehealth practices into its health system with some hospital-based virtual services performed exclusively in the private sector.^6^ Unfortunately, the legal infrastructure supporting the development of Telehealth systems was very weak. The Law Decision No. 1/227 governing the introduction of electronic and digital services in Lebanon issued in 2013 was not implemented and official Telehealth practice guidelines (TPG) and standards were still missing.^7^ The national E-Health Program foreseeing the incorporation of Telehealth and mobile health in respect to the principles of e-Health ethics (confidentiality, credibility, and privacy of health information) was not yet fully integrated into the health system.^7^ However, TPG and national policies were found to be essential for the proper implementation and assessment of remote services and for defining Telehealth quality metrics and performance measurement.^8^

As such, and in low-to-middle-income countries like Lebanon, Telehealth experience during COVID-19 appeared to be one of a kind. In Lebanon, the pandemic event coincided with the occurrence of a severe economic crisis and a massive explosion at Beirut port. Consequently, the health system and the technological infrastructure in the capital were severely destructed and hit simultaneously by liquidity crisis, bank withdrawal restrictions, lack of electricity, shortage in fuel, scarcity of medical equipment, hyperinflation and job losses.^9^ Under these circumstances, the financial crisis and the lack of available resources constituted additional barriers facing the adoption of advanced technology and the maturation and proper implementation of Telehealth.^10^ In addition, barriers associated with the socioeconomic status appeared to aggravate furthermore Telehealth inequities and disparities in the access to care during the pandemic.^11^

In this context, and equally to other health providers, dietitians were exposed to remote practices. In addition to the decrease in immunity and the exacerbation of chronic diseases and malnutrition noted elsewhere, the food shortages and food insecurity caused by the socio-economic crisis exposed the Lebanese population at a greater risk of malnutrition aggravating more and more the health impact of COVID-19.^12^

To date, studies conducted on Telehealth use among dietitians in the Arab world during the pandemic have focused on the change in the workload, caseload, tasks and responsibilities of hospital dietitians, and the challenges faced in the nutrition management of COVID-19 patients.^13^ The use of social/mass media by dietitians to perform Telehealth during COVID-19 was explored in Arab countries (such as in the UAE, KSA, Jordan…) and in Lebanon.^14^ Telehealth uptake by Lebanese physicians and Telehealth application for mental health amid the pandemic were studied, and findings on the increase use of Telehealth via video consultations for diagnosis and treatment have been reported.^6,15,16^ Even though COVID-19 has imposed the rapid adoption of Telehealth, the performance of healthcare providers during telehealth sessions was positively impacted by the existence of legal frameworks and technological infrastructures.^17^ Thus, and since the Lebanese laws failed to determine in which practice Telehealth can be eligible and applicable, in the absence of TPG, Lebanese physicians were urged to apply the general provisions related to their responsibility, liability, in-clinic medical errors and misdiagnosis to the virtual setting in order to properly conduct Telehealth services ^7^.

Based on these observations, we may conclude that, although COVID-19 had caused a global surge in Telehealth uptake, there is a deep gap in the literature concerning Telehealth use in the absence of TPG, and in countries suffering from economic crisis and weak infrastructures. On the other hand, Telehealth modalities adopted by healthcare providers, and specifically by dietitians in the MENA region, to respond to the dynamic change induced by COVID-19 in the healthcare system are not well explored. Further, there is a limited evidence on the importance of Telehealth services for nutrition care in the post-COVID-19 period, and for the health care system stability in this region.

To fill this gap, we addressed several research questions by conducting an exploratory research study using a cross-sectional, anonymous online survey administered to LDs during the month of March, 2023. The survey explored Telehealth uptake among LDs during COVID-19 and the sustainability of its application beyond the pandemic. In addition, it identified Telehealth modalities, challenges, benefits and perspectives experienced by LDs in the absence of TPG and where poor infrastructure and economic crisis prevail.

To achieve this goal, this study addresses the following questions: (1) Did the use of Telehealth by LDs change at the onset of the COVID-19 pandemic? (2) What tools and modalities did LDs use during Telehealth consultations and in the absence of TPG? (3) What were the benefits and barriers facing Telehealth use among LDs in Lebanon? and (4) What was the perspective of LDs on Telehealth experience during and post-pandemic in Lebanon?

Hence, the main contributions of this study include the following:

− Analysis on Telehealth use among LDs amid the pandemic and in the absence of TPG, with identification of perspectives, barriers, facilitators and benefits.
− Applicability of Telehealth for nutrition care in the Lebanese context.
− Identification of challenges facing the continuous improvement of Telehealth and its future implications in Lebanon along with state-of-the-art recommendations for its sustainability beyond COVID-19.
− Recommendations toward the incorporation of authorized Telehealth platforms into the Lebanese health system along with the development of Telehealth national policies, technical regulations, standards, protocol of care, and professional development.
− Promotion of Telehealth as a strategic and sustainable model to be incorporated into the normal dietetic practice in countries suffering from conflicts and economic instability.

Finally, this study constitutes a cornerstone for the developing of the Lebanese Telehealth nutrition system and its national legal framework, policies and TPG. This will assure Telehealth proper implementation and sustainability beyond the pandemic event and the socio-economic crisis and contribute to the incorporation of a remote evidence-based nutrition care into the routine practice of the LDs.

## Methods

### Survey development and design

A cross-sectional study was performed using an online questionnaire adapted from a peer-reviewed questionnaire published previously in the litterature.^18^ The original questionnaire was modified by removing questions that were not aligned with the practices in Lebanon (such as the estimated percentage of the patient population covered by Medicare). The study questionnaire was divided into 4 sections with a total of 44 questions covering LDs’ demographic, educational and professional characteristics, Telehealth experience before and during the pandemic, tools used, barriers, facilitators and benefits, and LDs’ perspectives on their Telehealth experience. The questionnaire was pilot-tested and the pilot-testing feedback was used to develop the questionnaire final version.

The inclusion criteria of the study were: Lebanese dietitian, holder of a bachelor’s degree in Human Nutrition and Dietetics, licensed by the Lebanese Ministry of Public Health (MOPH), and practicing in Lebanon during COVID-19.

### Data collection

To maximize the suvey reach in the absence of a full list of LDs, and to avoid the risk of not receiving direct emails, a link to the survey’s Google form was shared among LDs through the Lebanese Order of Dietitians, social media platforms (WhatsApp, LinkedIn, and Facebook) and with nutrition faculties and alumni of Lebanese universities. To avoid overlap and duplication in data collection, LDs who previously completed the questionnaire were asked not to participate. Google Forms was used to collect the survey data.

### Ethical considerations

The study was reviewed and approved by the Lebanese American University Institutional Review Board (LAU IRB): LAU.SAS.RR1.29/Nov/2022. Informed consent was implied by voluntary completing the questionnaire. Google Forms settings were adjusted to ensure that personal data, such as emails and IP address were not collected. The participation in the study was anonymous, voluntary, and non-payable.

### Statistical analysis

The survey data were descriptively managed and analyzed using Statistical Package for the Social Sciences (SPSS, Version 28). The means and standard deviations (SD) were presented for continuous variables, whereas counts and percentages were presented for categorical variables.

## Results

### Participants’ characteristics

In total, 94 LDs were included in this study. The sample consisted mostly of females (98.9%) and young dieticians (mean (SD) age: 30.54 (6.41) years), with a master’s degree (58.5%), and a mean (SD) of 7.89 (5.7) years of experience in dietetics practice. Most respondents (87.2%) identified clinical nutrition as their primary practice area, concentrating on weight management (84%), food and nutrition consultation (58.5%), and diabetes care (45.7%). Table 1 describes the socio-demographic characteristics of the participants.

**Table 1.**
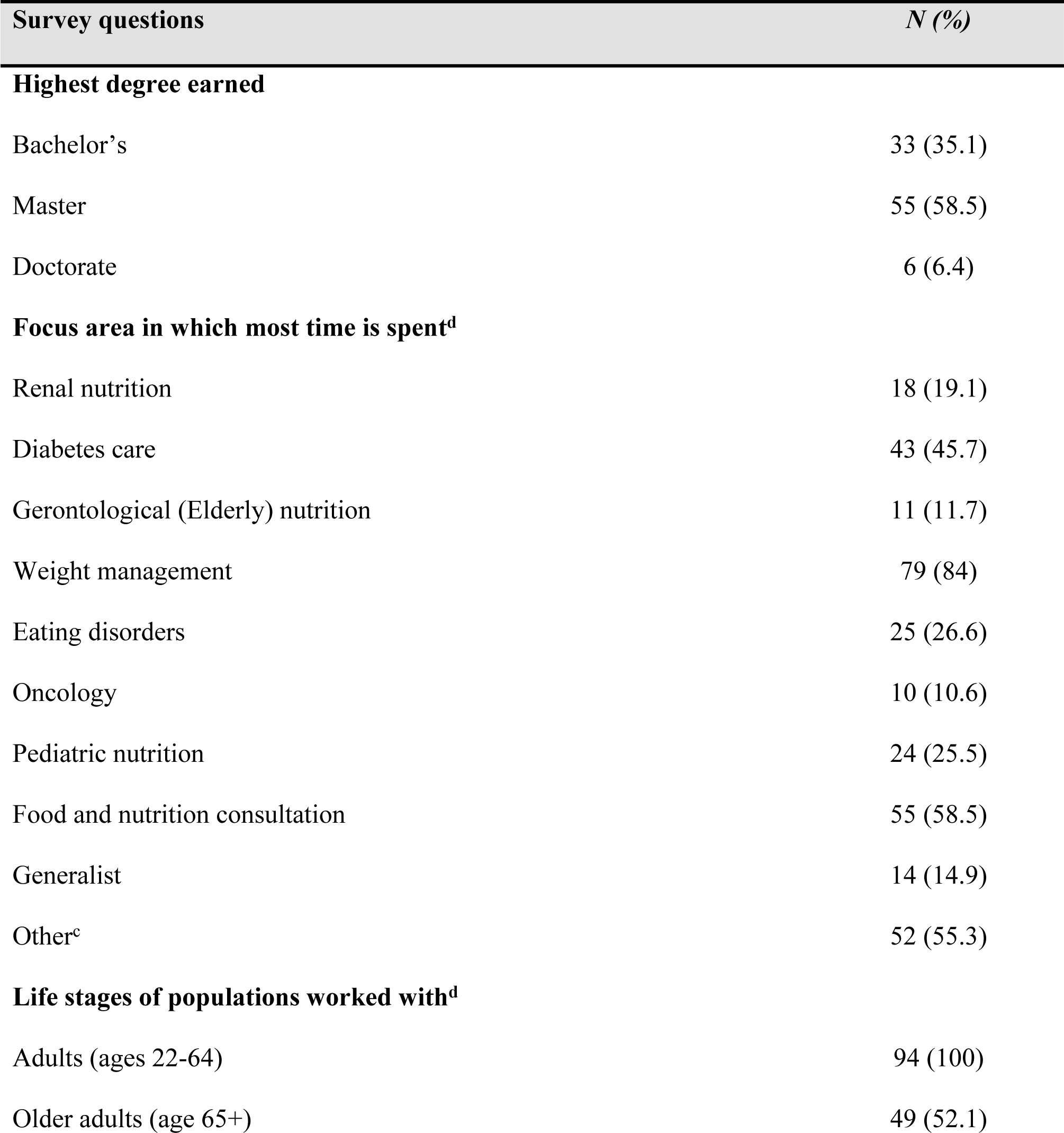
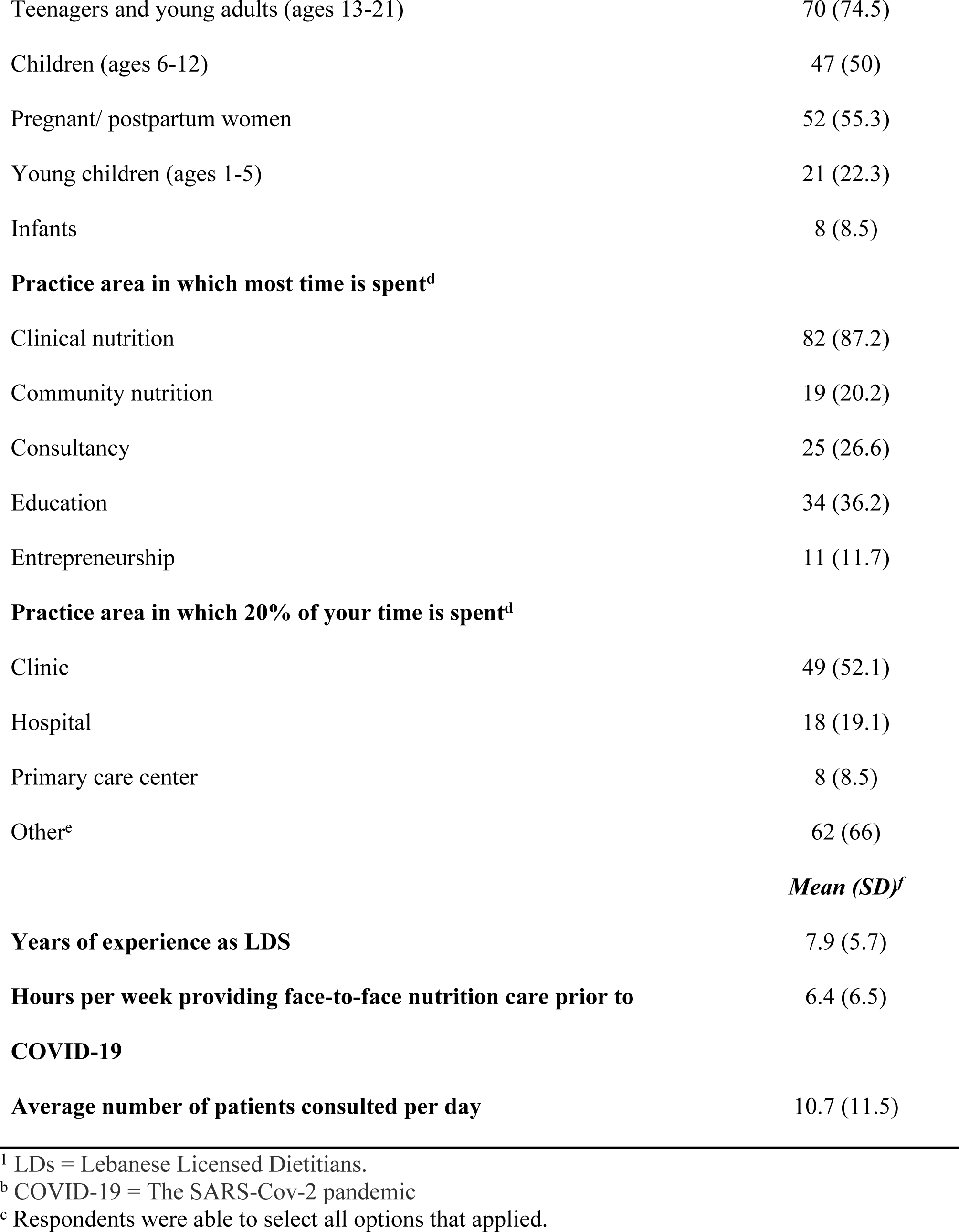
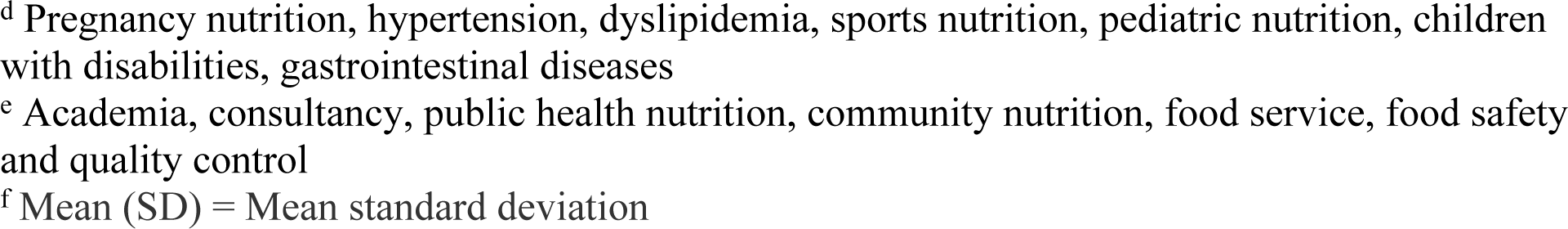
The Socio-Demographic Characteristics of a Cross-Sectional Sample of LDs^a^ Practicing in Lebanon and Using Telehealth During COVID-19^b^ (N=94)

### Telehealth experience during COVID-19 pandemic

The experience of LDs on Telehealth use for nutrition care prior to and during the pandemic are provided in Table 2. Although 48.4% (N=45) of the respondents reported using Telehealth for nutrition care before the pandemic, this proportion increased drastically to 97.8% (N=91) during COVID-19. LDs who reported using Telehealth before the onset of the pandemic had a mean (SD) of 1.43 (2.5) years of experience doing so. During the pandemic, the main tools used to provide Telehealth nutrition care and illustrated in Figure 1, were WhatsApp (90.3%), Zoom (72.0%), E-mails (41.9%), and telephone (39.8%). During Telehealth sessions, LDs relied mostly on video calls (83.9%) followed by voice calls (67.7%) and voice messages (65.6%) and the majority (51.6%) spent more than 30 minutes in direct contact with individuals or group to collect information on food and nutrition-related history (94.6%), patient history (89.2%), patient behavior (74.2%) and, knowledge, beliefs, and attitudes (72.0%). LDs communicated the results of Telehealth sessions to the referring medical provider by relying the most on social media applications (40.9%), e-mail (21.5%) and electronic medical record (20.4%).

**Figure 1:**
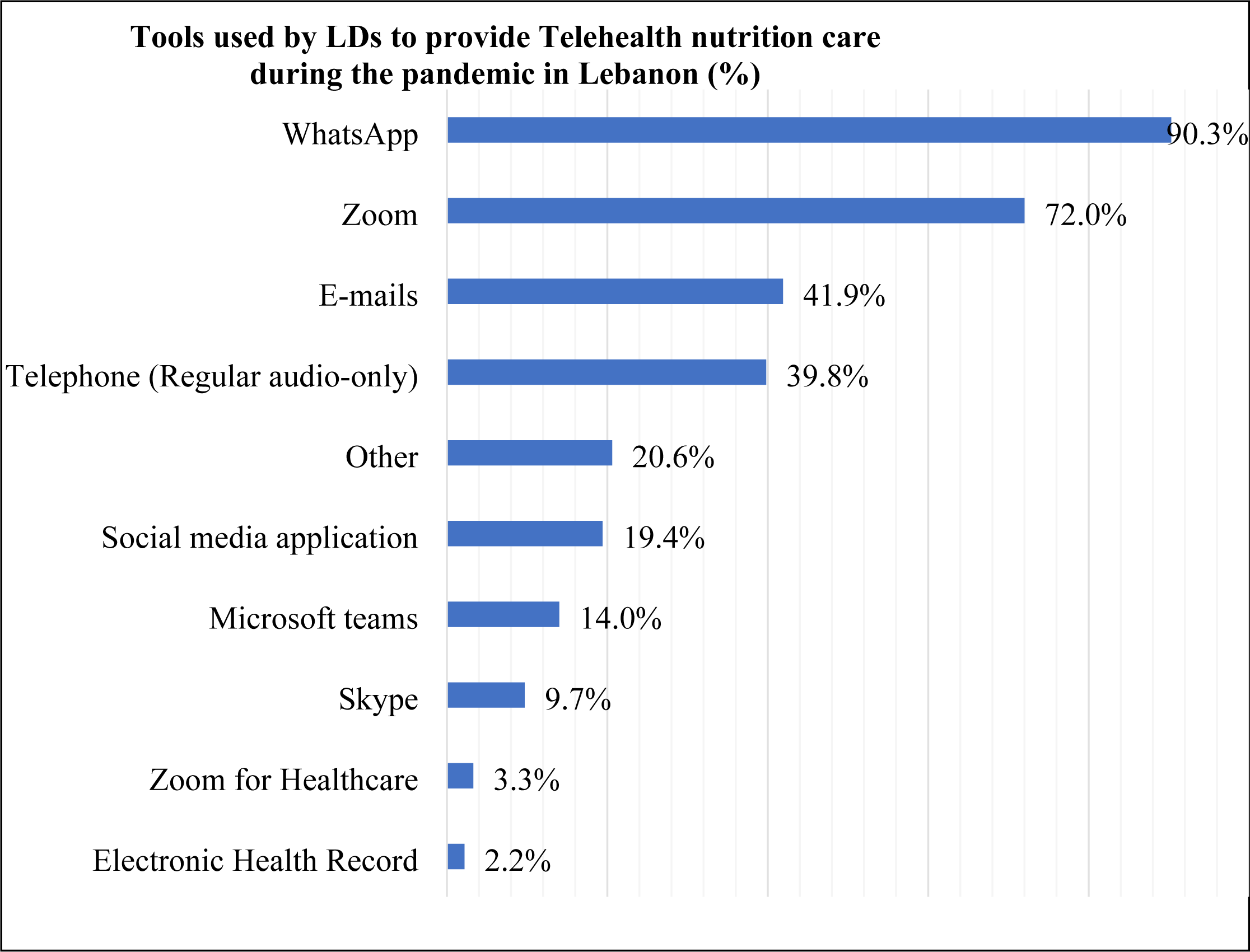
Tools Used by LDs^a^ to Provide Telehealth Nutrition Care During the Pandemic (%) (N=93)^b^. This figure shows the tools used by LDs to perform Telehealth for nutrition care during the pandemic. As reported, 90.3% of LDs used WhatsApp as their main tool, followed by Zoom applications, emails and Telephone However. Zoom for Healthcare and the Electronic Health Record were barely used. ^a^ LDs = Lebanese Licensed Dietitians ^b^ Respondents were able to select all options that applied

**Table 2.**
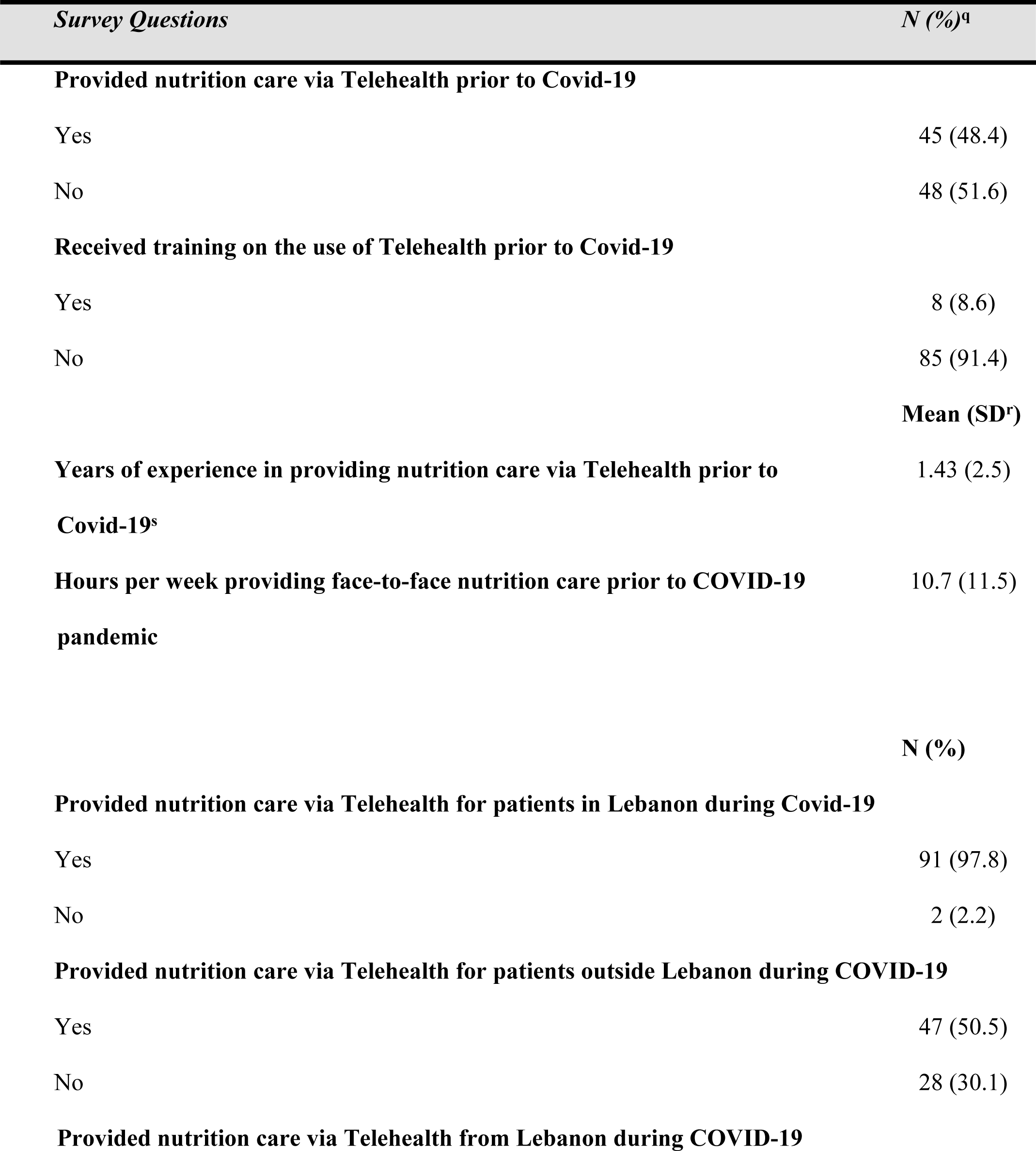
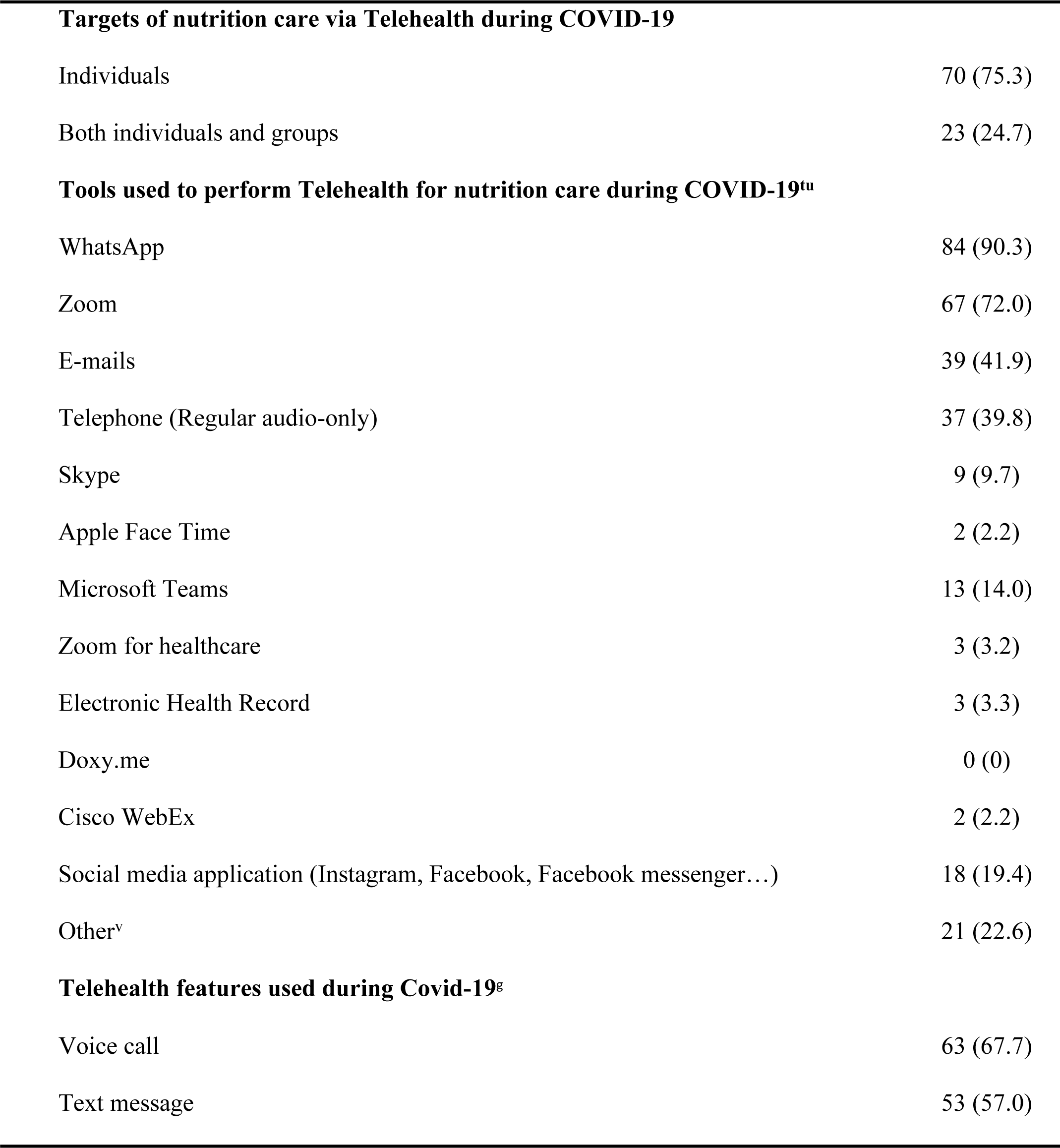
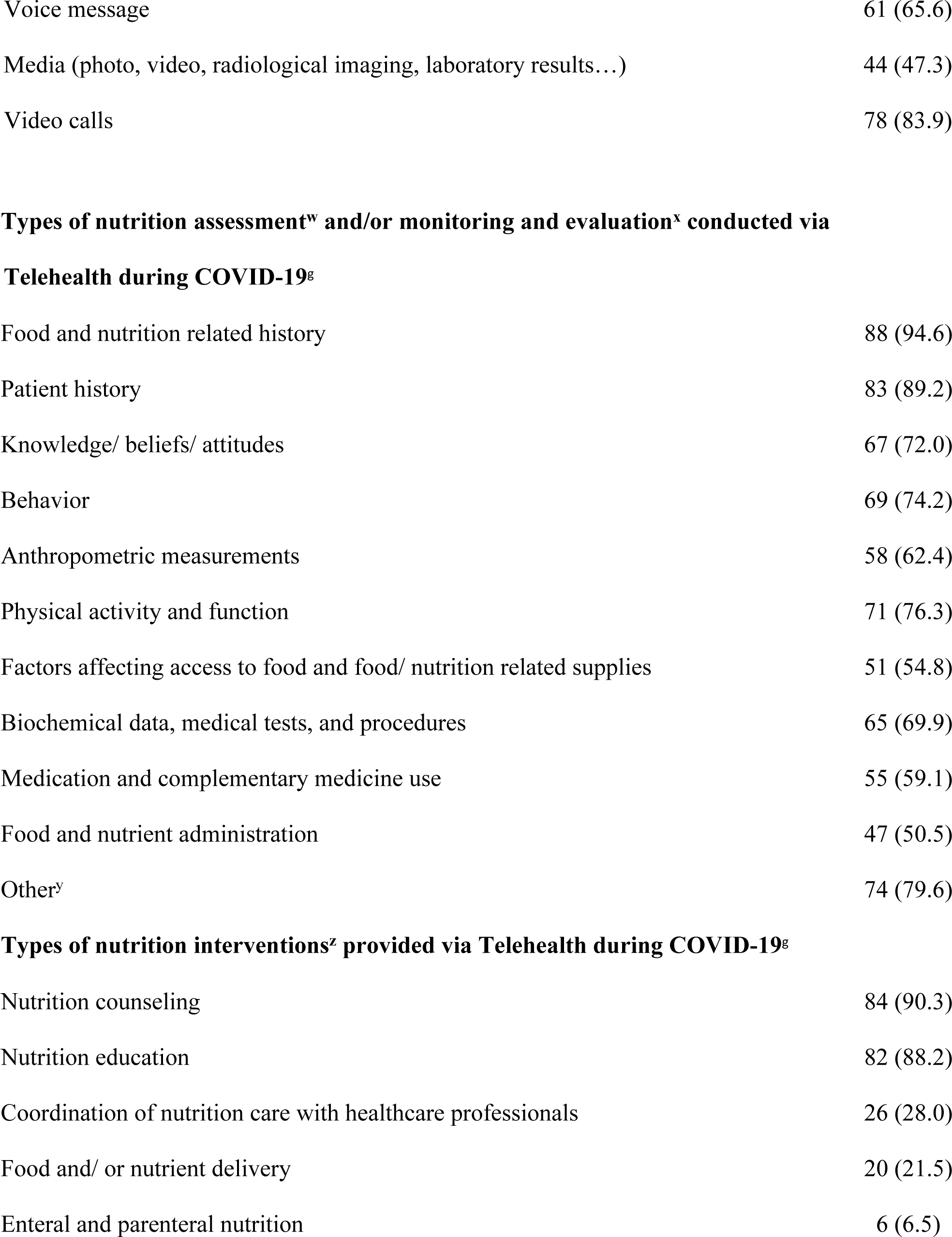
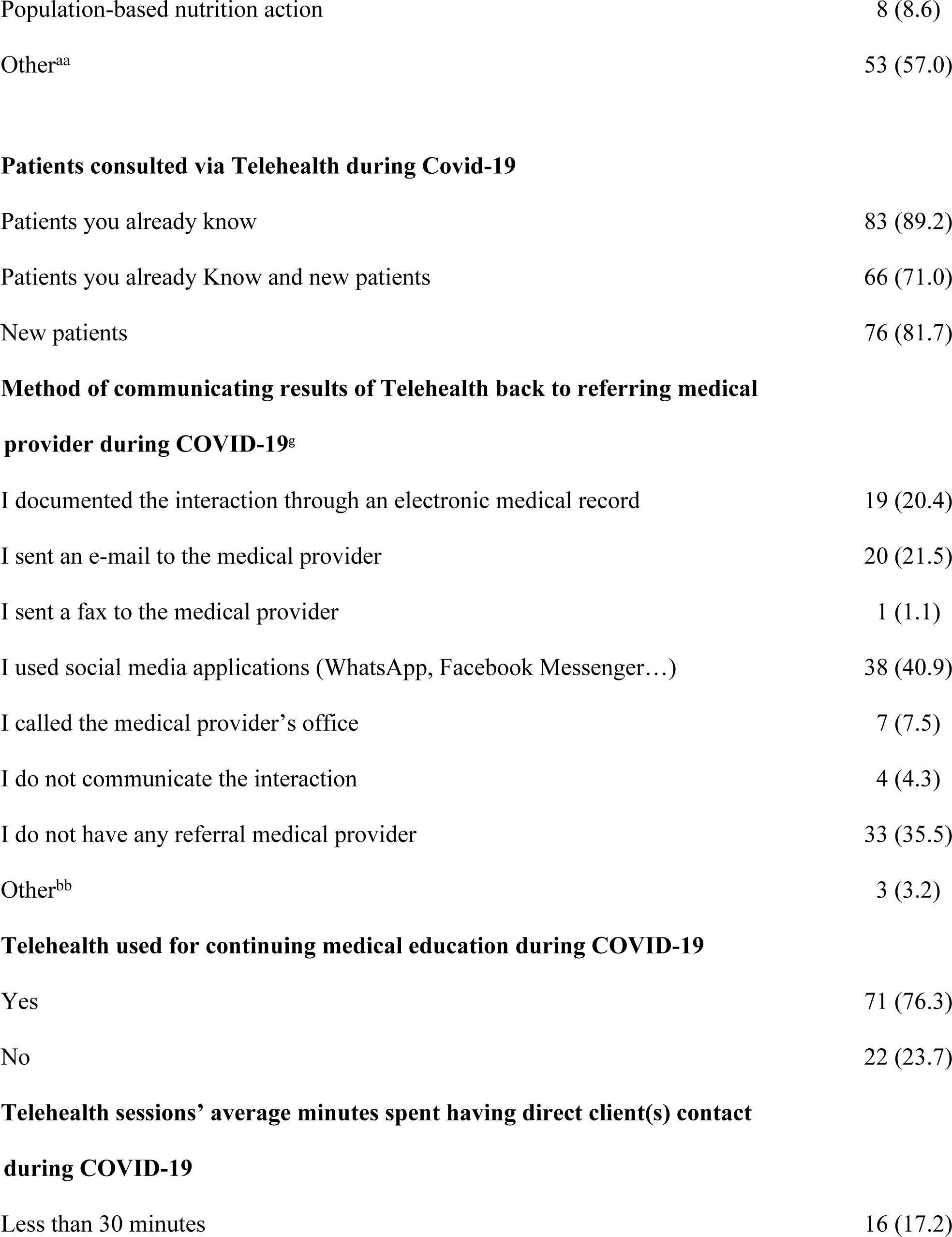
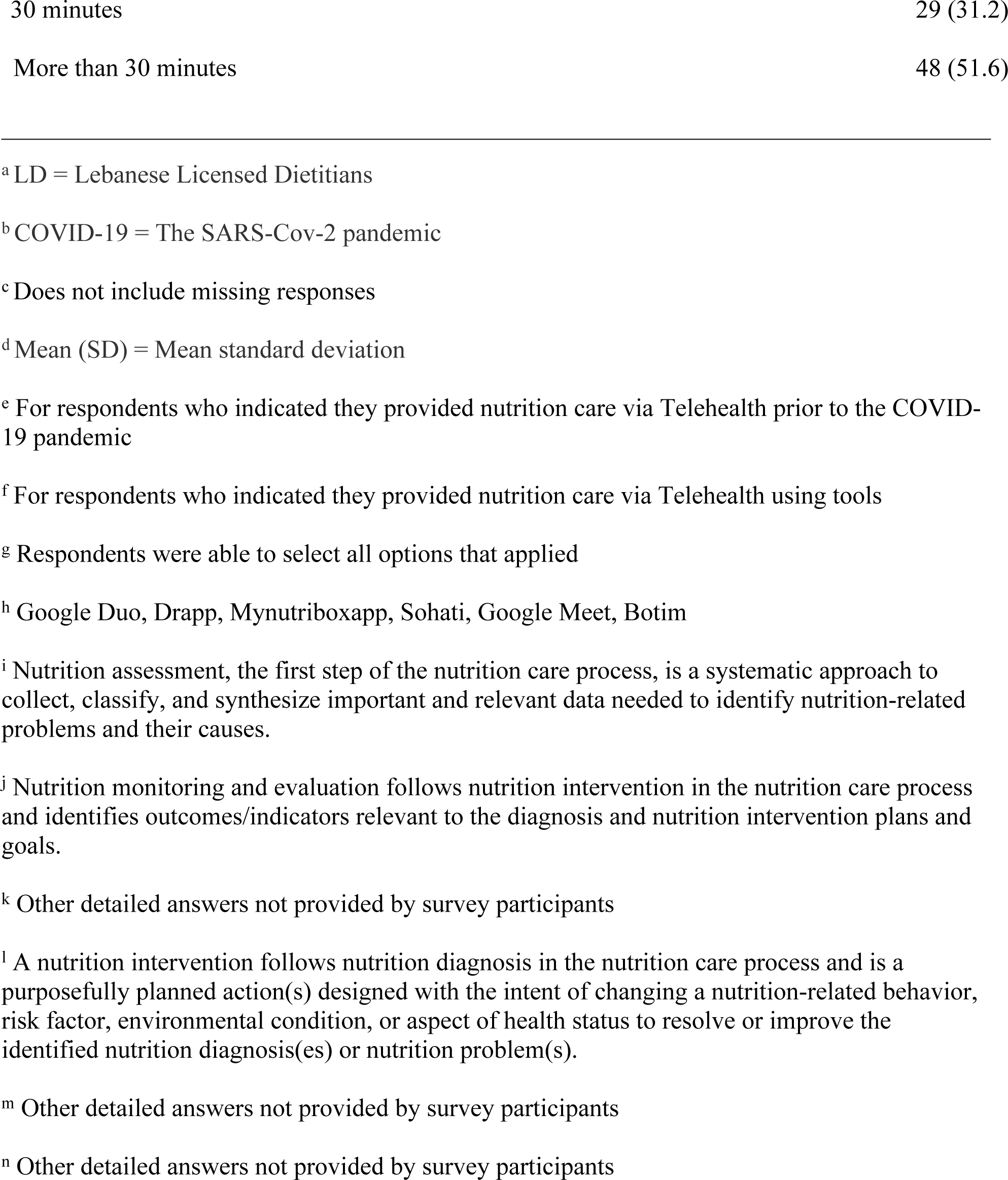
LDs° Respondents’ Experience in Providing Telehealth for Nutrition Care Prior to and During COVID-19^p^ in Lebanon (N=93)

### Barriers, facilitators, benefits and perspectives related to Telehealth use

The reported barriers, facilitators and benefits of Telehealth use during COVID-19 are illustrated in Table 3. The most reported barriers facing Telehealth nutrition care were bad connections (74.2%) followed by patients preferring face-to-face consultation (61.3%), and patients lacking technical literacy (33.3%). Among the benefits, LDs reported scheduling flexibility (83.9%), the decrease in practice-related costs (77.4 %), the promotion of technology acceptance and penetration (69.9 %) in addition to the compliance with COVID-19 social distancing measures (53.8%). As demonstrated in Figure 2, during the pandemic, most respondents did not receive any trainings on how to properly conduct Telehealth (93.5%) and did not consult any specific guidance (94.6%). The majority agreed that Telehealth is needed in Lebanon (78.5%), 64.6% believed that Telehealth is applicable in the Lebanese context and a high number expressed their willingness to use Telehealth after the pandemic (81.7%). Adding to this, 78.5% reported their interest in receiving future trainings on Telehealth for nutrition care and 74.2% expressed their need for specific TPG to properly conduct remote nutrition care (74.2%).

**Figure 2:**
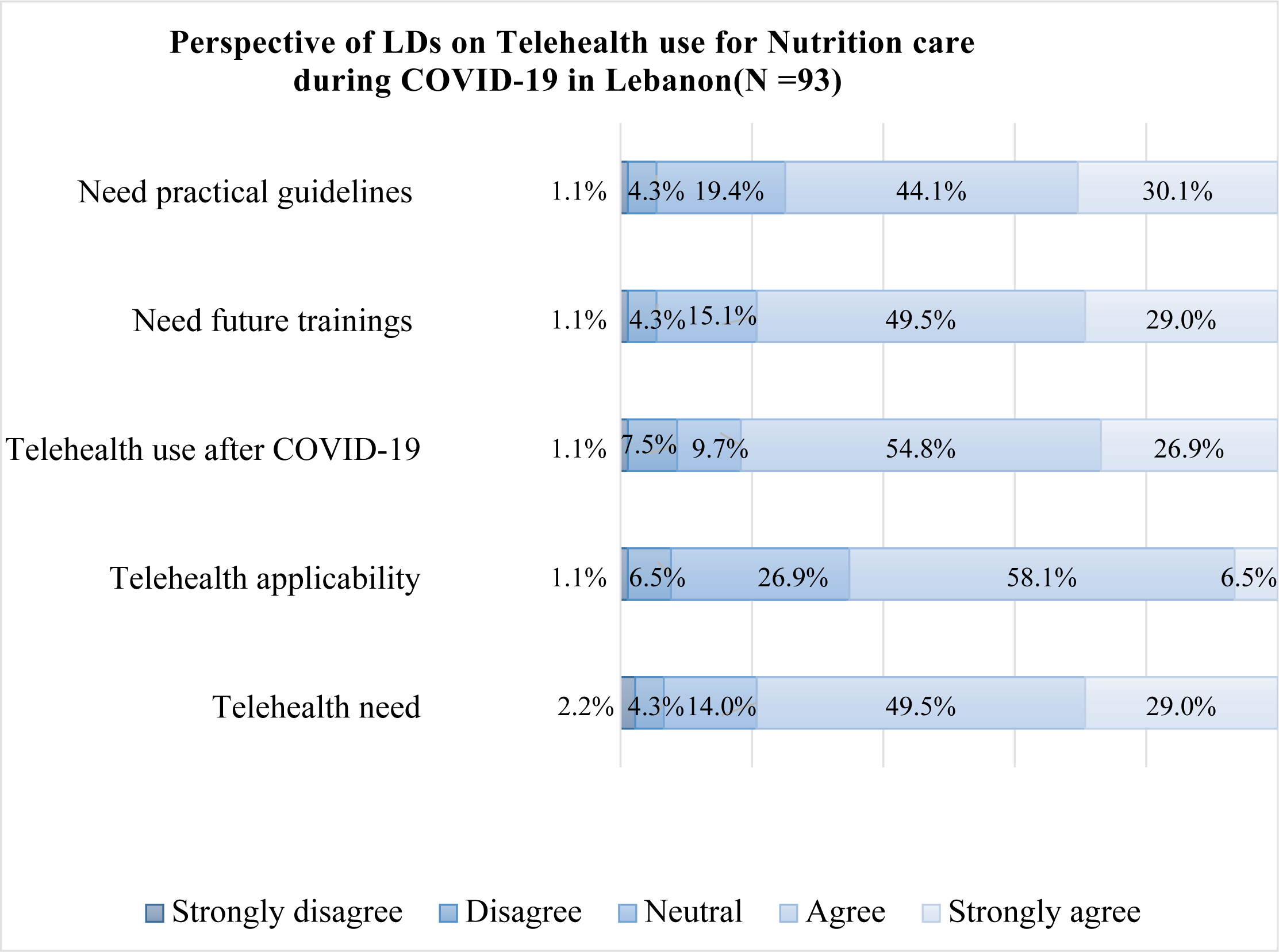
Perspective of LDs^a^ on Telehealth Use for Nutrition Care during The Pandemic (N=93)^b^ This figure illustrates the perspectives of Lebanese Licensed Dietitians (LDs) on telehealth use for nutrition care during the COVID-19 pandemic in Lebanon Using the Likert Scale (Strongly disagree. Disagree. Neutral. Agree, and Strongly agree), LDs responded on their need for practical guidelines, their need for future trainings, their willingness to use Telehealth post-pandemic in addition to their perspectives on Telehealth need and Telehealth applicability in the Lebanese context LDs responses are presented by percentage. ^a^ LDs = Lebanese Licensed Dietitians ^b^ Include missing responses

**Table 3.**
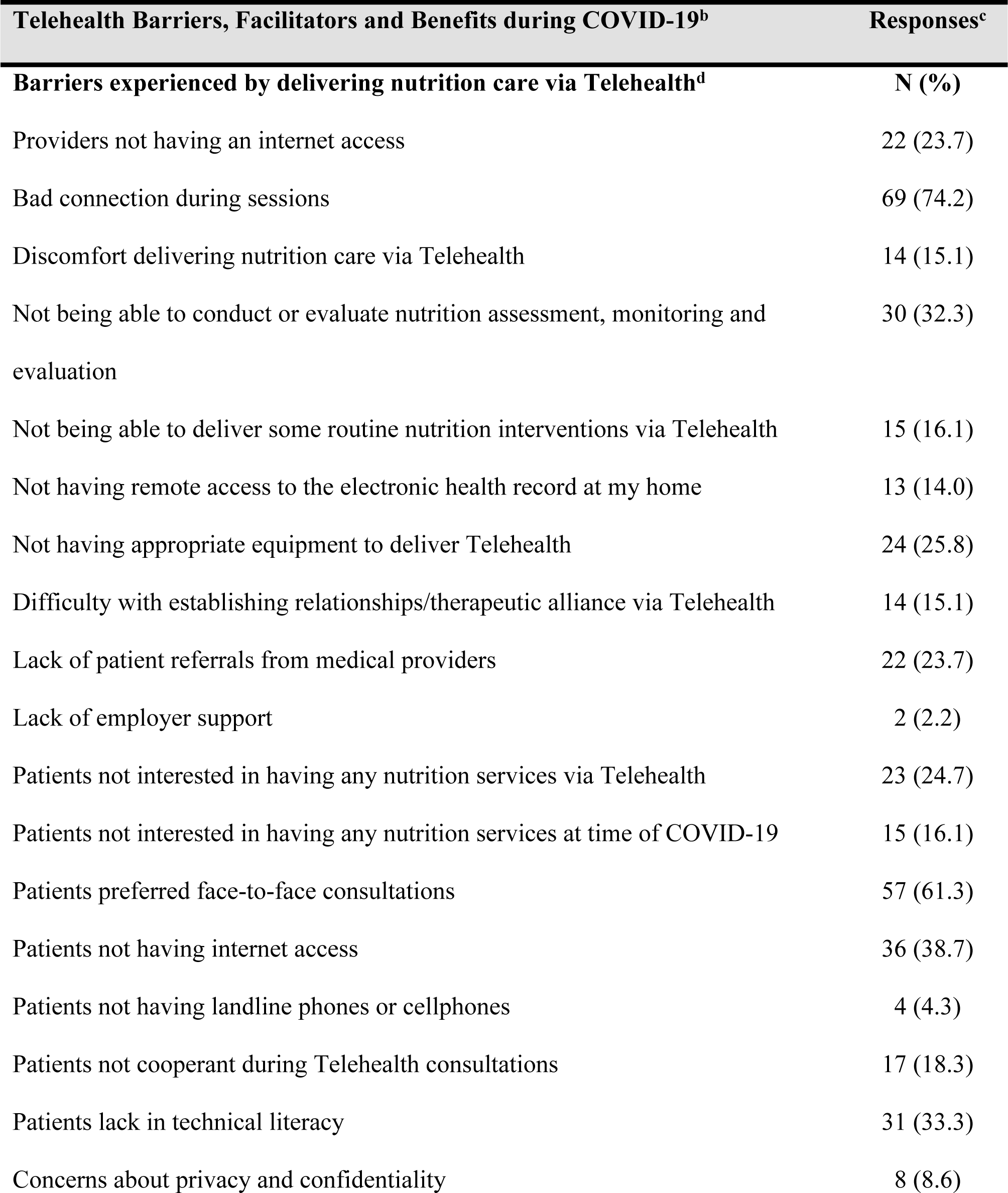
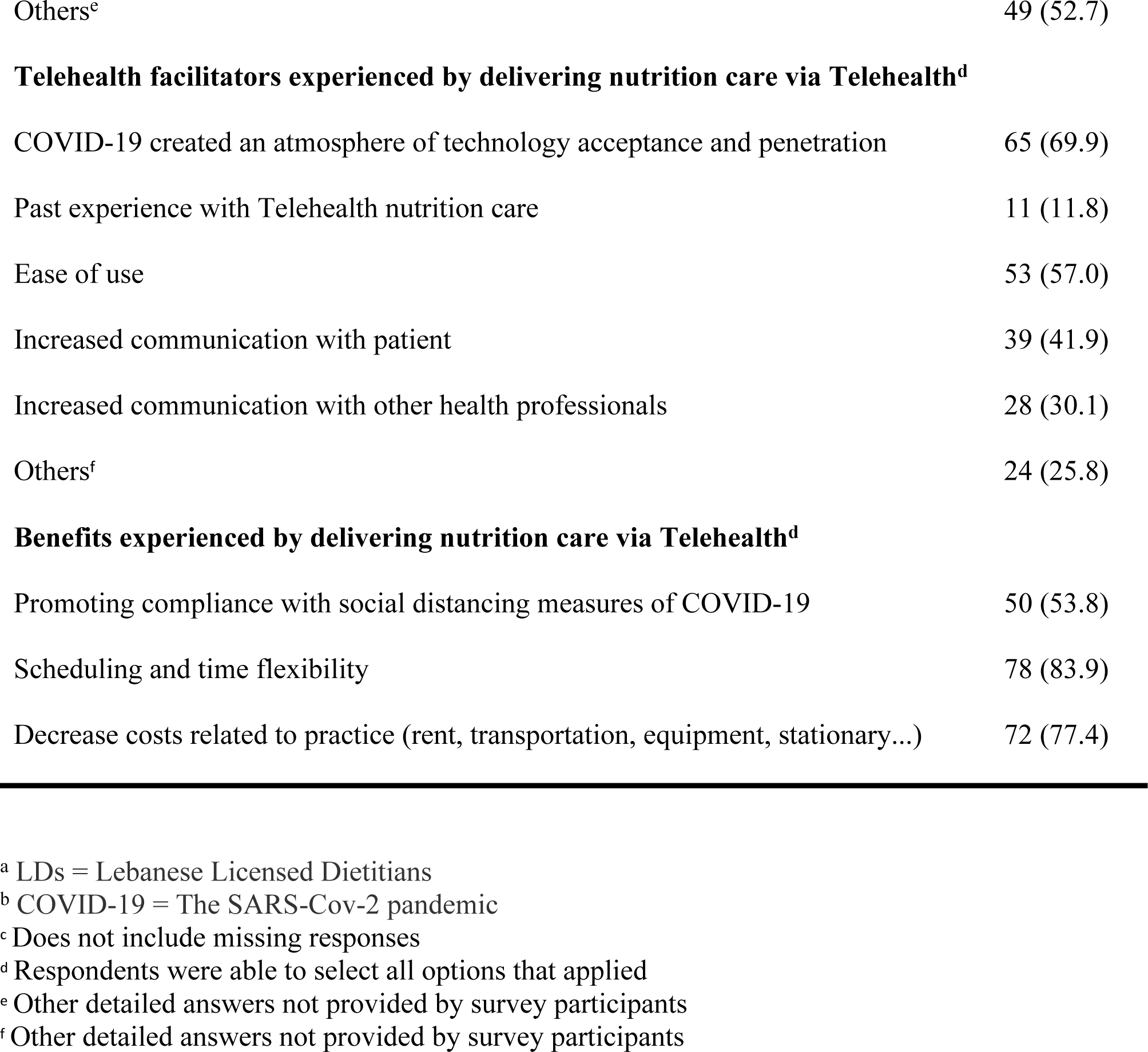
LDs^a^ Reported Barriers, Facilitators and Benefits of Telehealth Use during COVID-19^b^ in Lebanon (N=93)

## Discussion

This study is the first to investigate Telehealth use among LDs prior and during COVID-19 in the absence of national Telehealth practice guidelines (TPG), standards and best practices assuring ethical and safe practices. It identifies Telehealth tools, benefits and barriers, and offers insights on Telehealth applicability in the Lebanese context and its effectiveness and future sustainability.

Amid COVID-19, this study reports an increase in Telehealth use among LDs which is expected as it mimics the trend reported by other healthcare professionals in Lebanon, and by dietitians in other countries.^6,14,19^ Remarkably, the majority of LDs (97.8%) reported using Telehealth for nutrition care amid COVID-19. In time of pandemic, Telehealth is perceived as a viable tool able to overpass lockdown measures, assure patient safety, maintain the cycle of life, and improve the mental health and quality of life.^18,20^ However, we note a high use of Telehealth practices among LDs before COVID-19 (48.4%) whereas only 15% of dietitians in Italy and 37% of dietitians in the US performed Telehealth activities prior the pandemic.^20^ This could be related to the fact that some Telehealth activities were initiated in the country before the pandemic onset as alternative practices to encompass the impact of the socioeconomic crisis, notably the scarcity of resources, lack of job opportunities, and limitation of transportation.^6,21^ As such, and with limited financial resources, LDs might have resorted to remote modalities of nutrition care. More, and despite the pre-pandemic Telehealth activities, the majority concentrated on face-to-face consultations in clinics (82%) and hospitals, suggesting herein that Telehealth pre-pandemic practices were used to follow-up on LDs’ existing patients and/or for continuous education. Similar pre-pandemic observations were noted among Lebanese physicians who, with limited financial resources caused by the economic crisis, couldn’t travel to attend international conference and relied mainly on Telehealth for continuing medical education.^6^ More, the use of video-conferencing for Telehealth consultations was reported low among Lebanese physicians before the pandemic.^6^

In contrast, COVID-19 imposed the rapid shift to audiovisuals and video-conferencing. LDs embraced Telehealth with uptake being centered in private practices and among newly graduated LDs. The rapid uptake of Telehealth observed in the country could be explained by the fact that LDs population was young (mean (SD) age: 30.54 (6.41) years), familiar to new technologies and in priory exposed to smart applications, which came similar to a study conducted on Italian dietitians.^20^ Same findings were reported in the MENA region.^13^

Thus, LDs initiated Telehealth amid COVID-19 by relying on WhatsApp (90.3%) as a main tool followed by Zoom (72.0%), Emails (41.9%) and phone calls (39.8%). In parallel, Lebanese physicians reported using WhatsApp (79%), phone calls (77%) and email (63%), in addition to Zoom and social media platforms (such as Facebook, YouTube and Instagram).^6^ Remarkably, and during COVID-19, the use of mobile applications/software as Telehealth tools was highly observed in low-to-middle income countries, with preferences for audio-visuals (such as WhatsApp, Zoom, Facebook Messenger and Skype).^22^ This was reflected in our study, and showed that LDs were able to select Telehealth tools adapted to the country’s economic status despite the absence of guidance in this direction. These tools offered LDs an atmosphere similar to clinics, allowing direct interactions with patients (inside and outside Lebanon), collection of data, feedback, and evaluation. In contrast, the use of Zoom for healthcare and electronic medical record was minimal (around 3%) and exclusively reported among LDs working in hospitals. In fact, Electronic health care records were incorporated in some major private hospitals before the pandemic but their use was limited to the context of some practices.^6^

More, Telehealth platforms available in Lebanon (such as TrakMD, SohatiDoc, Doctori, and Drapp) were not been authorized by MOPH and the Lebanese Order of Physicians.^23^ Due to these facts, Lebanese physicians and patients reported their preference in using WhatsApp instead of Telehealth platforms as it offered them spontaneous, informal and free of charge conversations.^23^

The findings of this study reflect the failure of the Lebanese nutrition care system in integrating Telehealth into the routine practice. As a result, LDs were not professionally equipped to initiate remote consultations and their profession and resilience were challenged by the pandemic. This can explain their resort to mobile and social media applications (such as WhatsApp, Facebook Messenger…) (40.9%) and the use of direct emails (21.5%) to communicate with referring medical providers. Social media applications emerged hereafter as alternative Telehealth tools offering direct communication with patients, medical providers and colleagues, and indirect communication with virtual profiles via its platforms. In contrast to other scientific-based applications and business communication platforms, social media gave LDs the opportunity to create dietetic contents enhanced with graphics and effects allowing the exponential dissemination of flow of knowledge with interdisciplinary integration. Hence, dietitians were able to go beyond counselling and nutrition care and to address concerns, social trends and common interests experienced in times of pandemic.^14^

Relying on this, this study acknowledges that, in the absence of authorized Telehealth systems and legal framework, and with the presence of weak infrastructure, socio-economic crisis and limited resources, resorting to social media during the pandemic supported the resilience of LDs and offered them business opportunities and marketing experience to grow. In the same direction, the use of social media was largely reported in other Arab countries (such as the UAE, KSA and Jordan) and appeared to be an effective Telehealth tool during COVID-19, despite concerns over privacy and data misuse.^13^

During the remote nutrition care, LDs identified several barriers centered around bad connections (74.2%), patients preferring face-to-face consultation (61.3%) and patients lacking technical literacy (33.3%). They expressed their discomfort during Telehealth sessions and faced barriers specific related to the dietetic profession such as difficulties in collecting physical data, and conducting nutrition assessment (32.3%). In addition, cultural barriers related to privacy concerns identified in our study came similar to those reported in the US and in Arab countries (such as Egypt, Jordan, the United Arab Emirates (UAE…).^13,18,24^

One of the notable findings to emphasize on in this study, is the delivery of nutrition care outside Lebanon which was reported by 50.5% of LDs. Such practices can have legal implications in normal circumstances and were only possible during the pandemic due to the relaxation of Telehealth regulations.^18^ Consequently, the lack of knowledge in the legal and ethical aspects of Telehealth use between countries, points out on the necessity of TPG and professional training. In similar, a minority of physicians in KSA reported being aware of the existent regulations governing Telehealth proper delivery via smart devices, which emphasizes on the need for guidance regulations.^25^

Nevertheless, LDs were able to identify Telehealth benefits such as scheduling flexibility (83.9%), decrease in practice-related costs (77.4%), and promoting compliance with social distancing measures (53.8%). The mean (SD) rating of their overall experience scored 6.62, on a scale from 1 to 10. They found Telehealth being effective and applicable in Lebanon and highly expressed their need for TPG and trainings. They were aware of the absence of legal framework and professional support assuring the protection of exercise, confidentiality, security, and privacy (Table 3). Similar needs were expressed by dietitians in Italy and in the UAE.^13,20^

In conclusion, the study results complement the findings noted among dietitians in other countries, and among other medical professionals in Lebanon. The Lebanese Telehealth experience appeared to resemble the experience described in the MENA region during COVID-19.^26^ Yet, we demonstrate that Telehealth is effective, applicable and needed in the Lebanese context as an alternative and resilient tool in time of pandemic, disaster and severe crisis with willingness to use it after COVID-19.

### Strengths and Limitations

This study pioneered in exploring Telehealth use among LDs in the absence of TPG during COVID-19. To provide meaningful results, enable comparisons and increase the survey’s reach, we adapted a questionnaire previously used in other studies to the Lebanese setting, and diffused an online survey via official channels and social media platforms. As such and in a short period of time, data was collected simultaneously from several participants without influencing any of their responses. However, the results were limited by being self-reported rather than observed, and the data collection might be subject to social desirability bias such as participants might have changed their answers in order to reflect a more professional practice. Further, the survey was shared among selected professional Facebook groups, which could lead to selection bias. Participants who have previously used Telehealth might have been pre-selected by the topic, i.e., being subject to self-selection. On the other hand, and as noted in other studies, the LDs population was relatively young with few years of experience in the dietetic practice.^18,20^ All these limitations might impact the generalizability of the findings for the larger LDs community. Finally, as this study was performed at the final stages of the pandemic, some answers might be subject to recall bias.

### Recommendations and future implications

Conducted in March 2023 at the final stages of the pandemic, this study might enrich the literature by exploring Telehealth use over multiple episodes of virus waves and lockdowns. By presenting the experience of a country suffering from a severe economic crisis that exacerbated furthermore the burden induced by the pandemic, this study was able to tackle Telehealth application during economic crisis and disaster events. Thus, it promotes Telehealth as a potential strategy of nutrition care to be incorporated into the normal practice in countries suffering from economic instability and conflicts like Lebanon.^22,27^

More, the benefits reported in this study present a rationale for telehealth sustainability and future implications beyond the pandemic in Lebanon. As demonstrated in the literature, the willingness to use Telehealth after the pandemic is reported high among those who used Telehealth before COVID-19^29^. This was reflected in our study and shows that LDs can play a major role in the sustainability of Telehealth practices in the near future. More, this study recognizes that Telehealth sustainability can offer low-to-middle income countries like Lebanon, the opportunity to achieve the Sustainable Development Goals (SDGs) of quality, accessibility and availability of health services to all.^28^ In this dimension, Telehealth sustainability and proper implementation could not be achieved without regulated and reliable Telehealth platforms able to secure personal and medical data, to assure patient’s privacy and confidentiality, and to protect the dietitian-patient relationship.^29^ Relaxed regulations during the pandemic have allowed the use of modalities such as social media and mobile applications, but health systems need to incorporate reliable platforms with appropriate privacy protection and validated security.^30^ Taking into account the weak infrastructure and the scarcity of resources in Lebanon, we suggest developing a unified Lebanese Telehealth nutrition platform for the public and private sector.

This correlates well with recommendations of other studies to use platforms equipped with interface for nutrition care such as the Academy of Nutrition and Dietetics Health Informatics Infrastructure (ANDHII) and to overcome problems related to interoperability between different health systems.^31, 32^ Developing TPG for nutrition care are fundamental for a safe and evidence-based standardized practice. These key elements can be translated into recommendations to the newly elected Lebanese Order of Dietitians to develop and harmonize, along with MOPH and other stakeholders, Telehealth legal framework comprising national policies, technical regulations, standards and protocol of care. These regulations must be complemented by the adoption of innovative technologies able to achieve Telehealth equity and accessibility among citizens and residents.

Beyond COVID-19, Telehealth sustainability requires the accreditation of Telehealth system for quality assurance, continuous improvement, evaluation of performance, standardization of protocols and operations, dissemination of Telehealth culture and acceptance, and protection of patient experience.^33^ Developing a competent and skilled Telehealth workforce within healthcare institutions is primordial and help to overpass any resistance facing the adoption of new technologies.^25,30^ Thus, we recommend continuous training and professional development for LDs (and other healthcare providers) on the proper use of Telehealth tools/modalities and on the ethical, privacy, legal and cultural aspects governing the virtual world.

The exceptional situation experienced during the pandemic catalyzed the integration of Telehealth activities within the health system and induced a paradigm shift in the practice and role of dietitians. At a glance, LDs were exposed to new models of nutrition care with innovative delivery modes from the virtual world. As a result, the sustainability of their carriers became closely linked to their capabilities and agility to embrace innovative practices and to converge their role with the emergent core technologies of the Fourth Industrial Revolution (Big data, Genomics, Artificial intelligence, etc.).^33^ More, personalized and precise nutrition care might replace the traditional treatment-oriented service in the healthcare system pushing hereafter dietitians toward the use of Telehealth nutrition platforms as part of their job routine.^2^ As such, and in the near future, innovation in the field of Telehealth and digital health for nutrition care are required in Lebanon.

## Conclusion

This study demonstrated the increase in Telehealth use among LDs amid COVID-19 and reports an overall satisfaction despite the absence of national regulations, TPG, and professional trainings. In this context, this study constitutes a foundation upon which Telehealth infrastructure and national TPG for nutrition care can be built upon. By presenting the experience of Lebanon, this study identified Telehealth as a strategic model applicable not only in time of pandemic and public health emergencies, but during economic crisis and scarcity of resources. Thus, it offers key elements for the future Telehealth sustainability and for the adoption of innovative patient-centered technologies. In this regard, further studies are needed in Lebanon to assess the effectiveness of Telehealth for nutrition care, its outcomes on the long run, the quality performance of its actors and the patient experience.

## Authors’ contributions

MA, MM, and RR conceptualized the study. MA, NC, and RR collected and analyzed the data. MA wrote the first draft with contributions from NC, MM, and RR. All authors reviewed and commented on subsequent drafts of the manuscript.

## Data Availability

If the data are all contained within the manuscript and/or Supporting Information files, enter the following: All relevant data are within the manuscript and its Supporting Information files.

## Acknowledgments

None

## Data Availability Statement

Generated or analyzed Data of this study are included in the supplementary information files of this manuscript.

## Research Snapshot

### Research questions

Did Telehealth use by Lebanese Licensed Dietitians (LDs) change amid COVID-19? What tools, barriers, benefits, and perspectives were reported in the absence of practical guidelines?

### Key findings

Telehealth use increased among surveyed 94 LDs from 48.4% before COVID-19 to 97.8% during the pandemic, mainly using WhatsApp, Zoom, and E-mails. Barriers included bad internet connection, patients preferring face-to-face consultation, and lacking technical literacy; benefits included scheduling and time flexibility, decrease in practice-related costs, and compliance with social distancing measures. LDs found Telehealth applicable in Lebanon and expressed their need for trainings and guidelines for proper implementation and sustainability.

